# Does time-incorporated admission severity improve stroke prognostication?

**DOI:** 10.1101/2025.11.08.25339812

**Authors:** Zewen Lu, Halvor Næss, Matthew Gittins, Amit K Kishore, Craig J Smith, Andy Vail

**Author notes:** Corresponding author: Zewen Lu, Jean McFarlane Building, University of Manchester, Oxford Road, Manchester M13 9PL. **Author contributions** Zewen Lu: Conceptualisation; Methodology; Study design and data analysis; Writing-original draft. Writing-review & editing. Halvor Næss: Data management and acquisition; Ethical approval; Writing-review & editing. Matthew Gittins, Amit K Kishore, Craig J Smith and Andy Vail: Conceptualisation; Methodology; Supervision; Writing-review & editing. **Conflicting interests** All authors declared no potential conflicts of interest concerning the research, authorship, and/or publication of this article. **Funding** None. **Ethical considerations** This study was approved by the Regional Ethics Committee for Medical and Health Research Ethics in Western Norway (approval no. 2012/1483 Norstroke). **Data Availability** All data produced in the present study are available upon reasonable request to the authors.

## Abstract

**Introduction:** The dynamic relationship between time since symptom onset and severity on admission is poorly incorporated in stroke prognostic models. We sought to develop a novel approach and compare it with alternatives.

**Methods:** We developed admission stroke severity centiles to contextualise a patient’s stroke severity relative to others admitted at a similar time post ictus. We conducted a secondary data analysis to compare four prognostic models, including the novel severity centiles and other time-incorporated approaches. Models were evaluated using Pseudo-R squared, Bayesian Information Criterion, and Area Under the Curve.

**Results:** Time since symptom onset not only predicts outcomes but also modifies the prognostic value of severity, with delayed admission amplifying its impact on functional dependency and mortality in acute ischaemic stroke (AIS) patients [Adjusted Odds Ratio (OR) (95% CI) = 1.02 (1.01 to 1.04); Adjusted OR (95% CI) = 1.03 (1.01 to 1.05), respectively]. Centile-based method offered a dynamic interpretation of stroke severity for both stroke types, demonstrating a prognostic signal comparable to traditional time-incorporated models. For AIS patients, each unit increase in centile corresponded to a 5% increase in the odds of functional dependency Adjusted OR (95% CI) = 1.05 (1.04 to 1.05) and a 5% increase in the odds of mortality Adjusted OR (95% CI) = 1.05 (1.04 to 1.07). Similar results were obtained for intracerebral haemorrhagic (ICH) patients. While covariate adjustment improved overall model performance, it did not alter the ranking of time-incorporated methods. Time-incorporated approaches did not enhance model performance for ICH patients, possibly due to a smaller sample.

**Conclusions:** We are the first to construct centiles for admission stroke severity. Severity centiles based on a reference population offer a graphical tool to support risk assessment and outcome prediction. While this approach provides clinically interpretable measures, our results suggest that explicitly incorporating time into prognostic models offers slightly better predictive performance. We recommend using statistical models that allow the effect of stroke severity to vary according to the time since symptom onset to assessment.

## Introduction

Stroke remains one of the leading causes of disability and mortality worldwide, with clinical outcomes intricately influenced by the rapid progression of neurological deficits that occur from symptom onset to hospital admission ^1,2,3^. The National Institutes of Health Stroke Scale (NIHSS) is the most widely used tool for assessing stroke severity in acute settings ^4^. Developed to standardise and quantify the extent of neurological impairment, the NIHSS evaluates a range of neurological functions, including consciousness, motor abilities, sensory perception, language, and coordination, through a comprehensive series of 15 items ^5^. Each item is scored individually, and the scores are summed to yield a total NIHSS score that ranges from 0 to 42, with higher scores indicating greater neurological impairment.

However, this measurement is only a single observation within a rapidly evolving process, where the severity at the time of assessment is inherently tied to the elapsed time since symptom onset ^6^. This relationship is complex and bidirectional; more severe symptoms may prompt earlier hospital presentation, while delays in admission can impact the progression of symptoms and subsequent stroke severity assessment. Despite this interdependency, existing prognostic models in stroke research often overlook the critical role of time from symptom onset to assessment and its interaction with admission severity, potentially limiting the efficiency of prognostic models ^7,8,9^. Our recent systematic review highlights this gap, showing a lack of time-incorporated models that consider these dynamics ^10^. This omission is particularly striking given that time from symptom onset is already recognised as a critical factor in stroke survival and recovery, as emphasised by the “FAST” campaign in the UK, highlighting the need to incorporate it more effectively into prognostic models ^11^.

In this study, we aimed to address this limitation by investigating whether time-incorporated admission stroke severity improves the performance of prognostic models in predicting functional outcomes and mortality. We introduced a novel approach by constructing admission severity centiles that account for the time from symptom onset ^12^. By assessing several methods of including time to assessment within a prognostic model of two key health outcomes, our research sought to offer a more comprehensive prognostic model that accounts for both the relative position in severity distribution and the elapsed time to assessment.

## Methods

### Study settings and participants

We conducted a secondary analysis of data from the Bergen NORSTROKE study ^6^, including patients with acute ischaemic stroke (AIS) or intracerebral haemorrhage (ICH) admitted to Haukeland University Hospital (2006–2021). Admission stroke severity was assessed using the NIHSS. Patients without recorded NIHSS scores, unknown symptom onset times, transient ischaemic attacks, unspecified stroke types, or symptom onset beyond 12 hours were excluded. The complete process of data cleaning and patient screening is shown in Appendix A, Figure 2.

### Centile-based Modelling

Admission stroke severity centiles, modelled using Generalised Additive Models for Location, Scale, and Shape (GAMLSS), provide a way to compare a patient’s stroke severity relative to others admitted at similar times post-symptom onset. We used a negative binomial distribution to fit admission NIHSS that allows the parameters of the distribution to vary as smooth functions of time from symptom onset to assessment ^12,13,14^. We modelled stroke severity centiles for AIS and ICH patients separately.

### Prognostic models and outcomes

We compared four prognostic models using binary logistic regression to predict functional dependency (modified Rankin Scale [mRS] >2) and mortality (mRS = 6) at day seven or discharge (if patients were discharged earlier), with various approaches to incorporate time since symptom onset to assessment. These included: (1) a model using only admission NIHSS without time, (2) independent covariates for NIHSS and time, (3) a model with time, NIHSS and their interaction, (4) a continuous centile model. Performance was evaluated using Nagelkerke’s R-squared, Area Under the Curve (AUC), and Bayesian Information Criterion (BIC). Admission NIHSS, time since symptom onset to assessment, and estimated centiles were used as explanatory variables, while outcomes were defined using dichotomised mRS scores as mentioned above ^15,16,17,18^.

We also performed multifactorial logistic regression, adjusting for the same confounders across four models. We used median imputation for missing age and medical history values, excluded patients with missing outcomes, and treated pre-stroke mRS as a three-level categorical variable (≤2, >2, and missing) without imputation.

## Results

### Admission stroke severity centiles

We summarised the patient characteristics of the study cohort in Appendix B, Table 3. We graphically illustrate the use of GAMLSS-modelled centiles to interpret stroke severity on admission, dynamically contextualising severity to aid clinical interpretation. In Figure 1 (a), For an AIS patient with an NIHSS score of six (black points), severity fell near the 60th percentile at one hour, meaning their stroke is more severe than 40% of similar patients. This increases to 70th at four hours and 80th at eight hours, showing rising relative severity with delayed admission. Figure 2(b) demonstrates a steeper centile shift for ICH patients.

**Figure 1.**
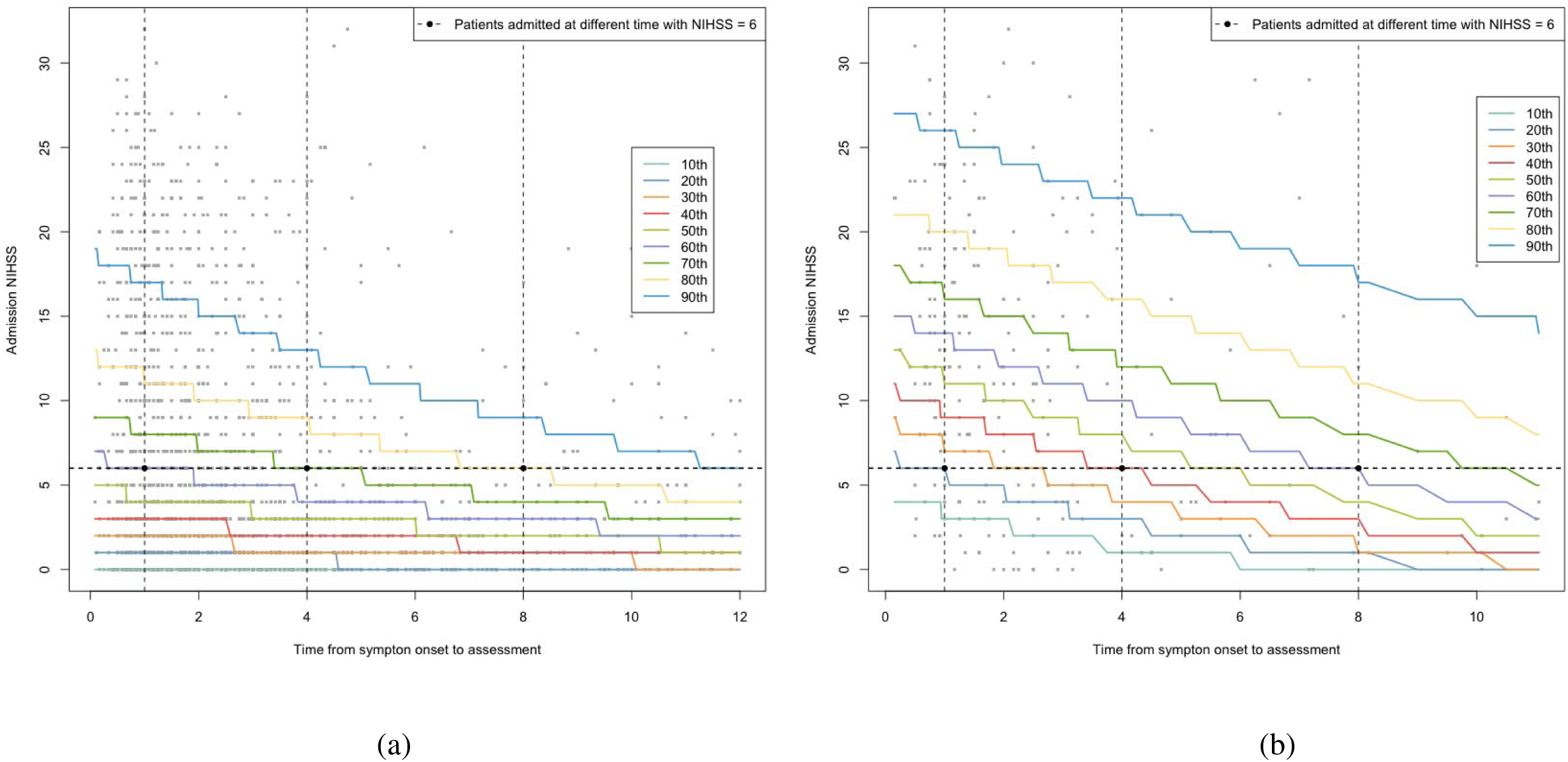
Stroke severity centiles modelled by GAMLSS for AIS patients (a) and ICH patients (b) in the study sample, respectively. From top to bottom, a symmetric group of centiles is plotted, including the 10th, 20th, 30th, 40th, 50th, 60th, 70th, 80th and 90th.

### Summary of explanatory variables

We summarised the results of the explanatory variables in predicting functional dependency in Table 1. For AIS patients, higher NIHSS scores at admission and a longer time to assessment are significantly associated with increased odds of functional dependency, as observed in Models 1 and 2. Model 3 highlights that the effect of severity on functional dependency can be modified by time: the effect increases with delayed admission odds ratio (OR) (95% confidence interval (CI)) = 1.03 (1.01 to 1.04) and adjusted OR (95% CI) = 1.02 (1.01 to 1.04). Model 4, which included continuous centiles as a measure of adjusted severity, showed that each unit increase in centile was associated with a 5% increase in the odds of functional dependency OR and adjusted OR (95% CI) = 1.05 (1.04 to 1.05). For ICH patients, a similar trend was observed in Model 1,2,3, though time to assessment was not statistically significant. Model 4 showed an increased risk of dependency with higher centile levels, still emphasising the predictive value of stroke severity as a relative measure when time is incorporated. Similar results in predicting mortality are summarised in Appendix C, Table 4.

**Table 1.** ORs with 95% CI and p-value of explanatory variables in predicting functional dependency.

| Models | Explanatory variable | Acute ischaemic stroke |  |  |  |  |  | Intracerebral haemorrhage |  |  |  |  |  |
| --- | --- | --- | --- | --- | --- | --- | --- | --- | --- | --- | --- | --- | --- |
|  |  | OR | 95% CI | P-values | AdjOR* | 95% CI | P-values | OR | 95% CI | P-values | AdjOR* | 95% CI | p-values |
| 1 | Admission NIHSS | 1.23 | (1.21-1.26) | < 0.01 | 1.24 | (1.21-1.27) | < 0.01 | 1.23 | (1.14-1.33) | < 0.01 | 1.27 | (1.16-1.38) | < 0.01 |
| 2 | Admission NIHSS | 1.24 | (1.21-1.27) | < 0.01 | 1.25 | (1.22-1.28) | < 0.01 | 1.22 | (1.13-1.32) | < 0.01 | 1.25 | (1.14-1.36) | < 0.01 |
|  | Time to assessment | 1.04 | (1.00-1.08) | 0.06 | 1.05 | (1.00-1.09) | 0.04 | 0.96 | (0.83-1.10) | 0.55 | 0.91 | (0.78-1.07) | 0.26 |
| 3 | Admission NIHSS | 1.17 | (1.13-1.21) | < 0.01 | 1.18 | (1.14-1.22) | < 0.01 | 1.13 | (1.01-1.26) | 0.03 | 1.15 | (1.02-1.30) | 0.03 |
|  | Time to assessment | 0.94 | (0.89-1.00) | 0.06 | 0.95 | (0.89-1.02) | 0.15 | 0.84 | (0.68-1.03) | 0.10 | 0.80 | (0.63-1.00) | 0.05 |
|  | Interaction | 1.03 | (1.01-1.04) | < 0.01 | 1.02 | (1.01-1.04) | < 0.01 | 1.04 | (0.99-1.09) | 0.11 | 1.04 | (0.99-1.10) | 0.13 |
| 4 | Continuous centiles | 1.05 | (1.04-1.05) | < 0.01 | 1.05 | (1.04-1.05) | < 0.01 | 1.04 | (1.03-1.06) | < 0.01 | 1.04 | (1.03-1.06) | < 0.01 |

\* We adjusted for age, sex, pre-stroke mRS, medical history including cerebral infarction, subarachnoid haemorrhage, myocardial infarction, angina, peripheral vascular disease, hypertension, diabetes, paroxysmal atrial fibrillation, chronic atrial fibrillation in the model

### Comparisons of model performances

The performance metrics for each model, including Pseudo-R², BIC, and AUC, are presented in Table 2, with ROC curves shown in Appendix H, Figures 3 and 4. Model 4, which incorporates time through centile-based severity measures, performed slightly less well than Models 1 to 3 across both AIS and ICH cohorts. For AIS patients, Model 4 showed marginally lower pseudo-R² (0.319 unadjusted; 0.506 adjusted) and AUC (0.788 unadjusted; 0.864 adjusted), along with slightly higher BIC values, suggesting a modest reduction in model fit and discrimination. A similar pattern was observed in ICH patients, where Model 4’s performance remained slightly below that of models incorporating time more explicitly. These findings suggest that while modelling severity via centiles offers clinical interpretability, explicitly including time (Model 3) in prognostic models may be more effective in enhancing predictive performance. Results were consistent in adjusted models and mortality predictions (Appendix D, Table 5 and Appendix H, Figure 5 and 6).

**Table 2.** Model comparisons: predicting functional dependency.

| Models | Acute ischaemic stroke |  |  |  |  |  | Intracerebral haemorrhage |  |  |  |  |  |
| --- | --- | --- | --- | --- | --- | --- | --- | --- | --- | --- | --- | --- |
|  | Unadjusted model |  |  | Adjusted model* |  |  | Unadjusted model |  |  | Adjusted model* |  |  |
|  | Pseudo- | BIC | AUC | Pseudo- | BIC | AUC | Pseudo- | BIC | AUC | Pseudo- | BIC | AUC |
|  | R<sup>2</sup> |  |  | R<sup>2</sup> |  |  | R<sup>2</sup> |  |  | R<sup>2</sup> |  |  |
| 1 | 0.326 | 2232 | 0.792 | 0.512 | 1913 | 0.868 | 0.299 | 199 | 0.824 | 0.452 | 235 | 0.885 |
| 2 | 0.327 | 2236 | 0.791 | 0.513 | 1916 | 0.868 | 0.301 | 205 | 0.827 | 0.458 | 240 | 0.887 |
| 3 | 0.337 | 2224 | 0.794 | 0.520 | 1907 | 0.869 | 0.318 | 207 | 0.833 | 0.470 | 243 | 0.895 |
| 4 | 0.319 | 2246 | 0.788 | 0.506 | 1926 | 0.864 | 0.240 | 211 | 0.793 | 0.389 | 249 | 0.857 |

\* We adjusted for age, sex, pre-stroke mRS, medical history including cerebral infarction, subarachnoid haemorrhage, myocardial infarction, angina, peripheral vascular disease, hypertension, diabetes, paroxysmal atrial fibrillation, chronic atrial fibrillation in the model.

## Discussion

Our findings demonstrated that appropriate inclusion for time-incorporated stroke severity can be an important component in good statistical performance in a stroke prognostic model. We are the first to develop the stroke severity centiles in a GAMLSS framework. Time from symptom onset to assessment is a critical predictor, particularly when evaluated in conjunction with severity.

The estimated continuous centile instead of the NIHSS scores alone may be more useful to better reflect the dynamic nature of symptom severity on admission. The stroke severity centiles constructed based on a reference population have the potential as a graphical tool that allows clinicians to assess a new patient’s risk and predict outcomes if the precise time is unavailable or uncertain.

The predictive ability of the centile method was comparable to models that explicitly included both severity and time, with no clear statistical advantage observed. The estimated continuous centile may provide a valuable but limited perspective that emphasises relative risk rather than the direct effects of severity and assessment time. For ICH patients, it is also because we cannot precisely estimate the centile with the small sample in this study. This can further lead to a poor model fit in the prognostic model. Future research will evaluate the performance of the centile method and other time-incorporated methods on a large ICH patient cohort.

As indicated in this study and the existing literature, the performance of prognostic models may vary depending on various outcomes, especially different ways of defining the favourable or unfavourable functional outcomes ^19,20^. We also assessed another common way of dichotomising mRS. From Appendix G, Table 8, the centile method did improve the model fit slightly only for AIS patients when mRS was dichotomised as 0 to 1 vs 2 to 6.

Dichotomising the outcome may reduce statistical power, and mRS dichotomisation is not recommended, particularly for analysing treatment effects in recent stroke trials ^21,22,23^. While unnecessary for developing prognostic models in observational studies, we still assessed time-adjustment methods using ordinal logistic regression. Treating mRS as its original scale could further improve model fit, but as shown in Appendix F, Table 7, results were consistent with binary logistic regression, partly due to the violation of the proportional odds assumption. This assumption, fundamental to ordinal logistic regression, requires that the effect of predictors remains constant across outcome levels. However, the Brant test (Appendix E, Table 6) indicated that the effect of the estimated continuous centile or other covariates (such as time and interaction) on the odds of moving to a higher category is not consistent across the levels of the ordinal outcome, showing the assumption was not met ^24^.

For ICH patients, time-incorporated methods showed no statistical improvement, as time to assessment was not a strong predictor. While these approaches capture severity changes over time, their added complexity may not enhance model fit if time-to-assessment lacks predictive value.

This study has several limitations. Our prognostic models are not intended for stroke outcome prediction but rather for evaluating how to incorporate time from symptom onset into prognostic models. We did not perform external or internal validation and analysed only short-term outcomes due to data availability. A major limitation is the sample size reduction caused by missing time data, which may affect the accuracy of time-incorporated centiles. Future studies with more complete time data are needed to validate these methods and address potential biases.

## Data Availability

All data produced in the present study are available upon reasonable request to the authors

## Supplementary Material

### Appendix A Data cleaning and patient screening

The complete process of data cleaning and patient screening is shown in Figure 2. The initial dataset comprised 6786 patients. Data cleaning excluded 3151 patients with unknown symptom onset and admission severity assessment times, 15 patients with missing admission stroke severity, and 76 with time measurement errors, resulting in 3544 patients. Screening further excluded 837 patients with NIHSS assessments over 12 hours from symptom onset, 389 patients with transient ischemic attacks (TIA), and one patient with an unknown stroke type. The remaining cohort consisted of 2044 patients with ischaemic stroke and 273 with intracerebral haemorrhage (ICH). After excluding those with missing modified Rankin Scale (mRS) outcomes, the final study cohort included 2040 ischaemic stroke patients and 272 ICH patients.

**Figure 2.**
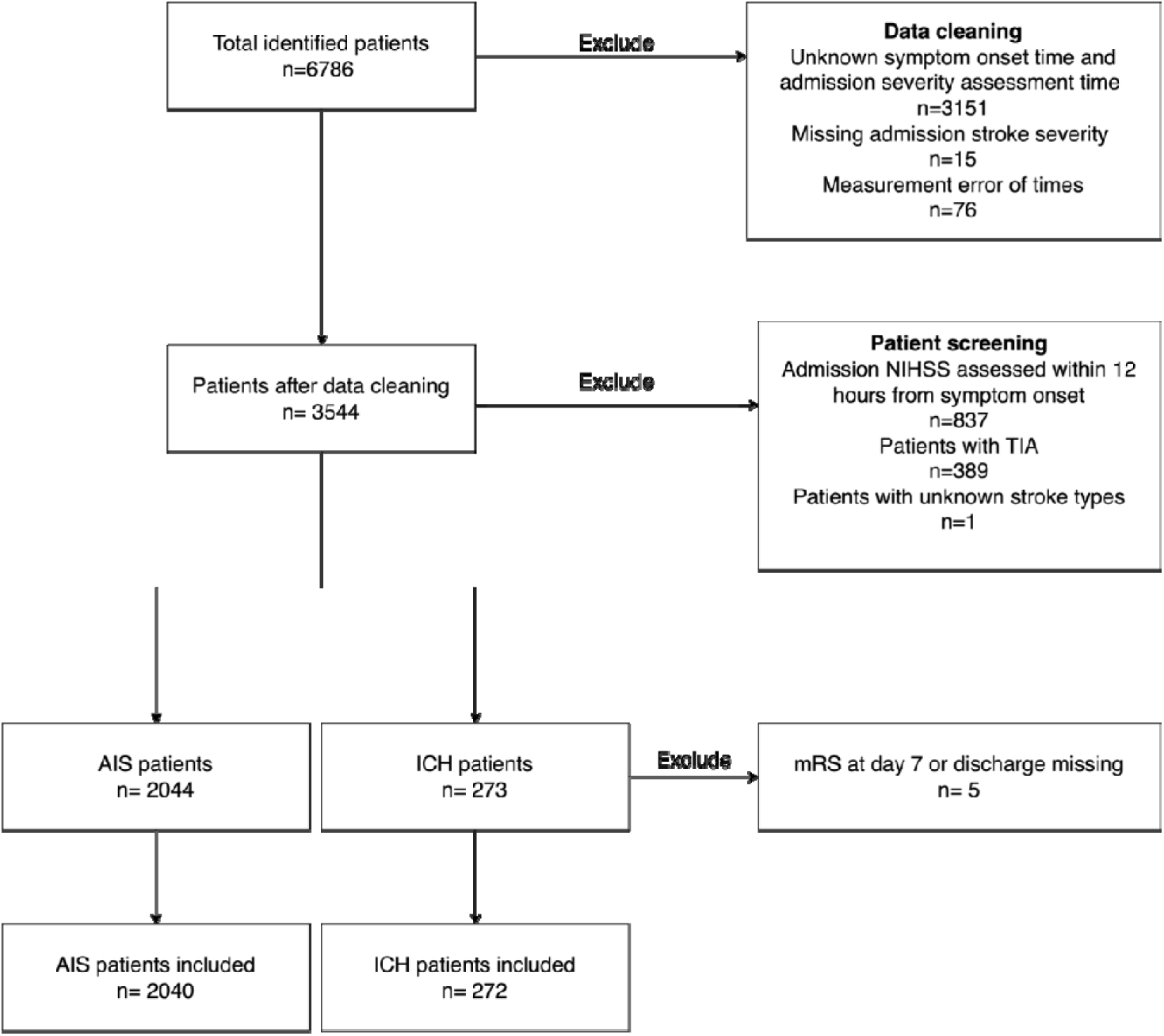
Flowchart of data cleaning and patient screening

### Appendix B Patient characteristics

We summarised the patient characteristics of the study cohort in Table 3. The median age was similar between the two groups: 74 years (IOR 62 to 82) for AIS patients and 75 years (IQR 65 to 81) for those with ICH. Males accounted for 43% of the AIS group and 41% of the ICH group. The time from symptom onset to NIHSS assessment was also comparable, with a median of 2 hours for both groups (IQR 1 to 4 for AIS and 1 to 3 for ICH). Median admission NIHSS scores indicated greater severity in ICH patients, with a score of 11 (IQR 4 to 8), compared to 3 (IQR 1 to 8) in the AIS group. Medical history was broadly similar between groups, including pre-stroke modified Rankin Scale (mRS) scores (median 0, IQR 0 to 1 for both), with minor differences observed in the prevalence of previous cerebral infarction (12% in AIS vs. 9% in ICH), subarachnoid haemorrhage (1% in AIS vs. 4% in ICH), heart attack (15% vs. 11%), and angina (14% vs. 11%). Other comorbidities, such as peripheral vascular disease, hypertension, diabetes, paroxysmal atrial fibrillation, and chronic atrial fibrillation, were similar across the groups.

**Table 3.** Patient characteristics.

| Characteristics | Total sample<br>n=2040 | Total sample<br>n=272 |
| --- | --- | --- |
| Age, median (IQR) yrs | 74 (62 to 82) | 75 (65 to 81) |
| Sex, (Male=1) % | 43 | 41 |
| Time from symptom onset to admission<br>NIHSS assessment, median (IQR) hrs | 2 (1 to 4) | 2 (1 to 3) |
| Admission NIHSS, median (IQR) | 3 (1 to 8) | 11 (4 to 18) |
| Pre-stroke mRS, Median (IQR) | 0 (0 to 1) | 0 (0 to 1) |
| Cerebral infarction % | 12 | 9 |
| Subarachnoid haemorrhage % | 1 | 4 |
| Myocardial infarction % | 15 | 11 |
| Angina % | 14 | 11 |
| Peripheral vascular disease % | 7 | 5 |
| Hypertension % | 52 | 52 |
| Diabetes % | 14 | 14 |
| Paroxysmal atrial fibrillation % | 11 | 11 |
| Chronic atrial fibrillation % | 9 | 10 |

### Appendix C Summary of explanatory variables in predicting mortality

As presented in Table 4, the explanatory variables when predicting mortality by day seven for AIS and ICH patients were assessed across the same four models. For AIS patients, higher admission NIHSS scores and shorter time to assessment increased the odds of mortality, as shown across Models 1, 2, and 3. Continuous centiles in Model 4 OR (95% CI) = 1.06 (1.04 to 1.07) and adjusted OR (95% CI) = 1.06 (1.04 to 1.08) also indicate an association with higher mortality, reflecting the relative severity at the time of assessment. For ICH patients, both absolute and relative severity measures are still critical in mortality risk stratification, though time incorporations have less effect for ICH patients.

**Table 4.** Summary of explanatory variables.

| Models | Explanatory variable | Acute ischaemic stroke |  |  |  |  |  | Intracerebral haemorrhage |  |  |  |  |  |
| --- | --- | --- | --- | --- | --- | --- | --- | --- | --- | --- | --- | --- | --- |
|  |  | OR | 95% CI | P-values | AdjOR* | 95% CI | P-values | OR | 95% CI | P-values | AdjOR* | 95% CI | P-values |
| 1 | Admission NIHSS | 1.17 | (1.13-1.20) | < 0.01 | 1.15 | (1.12-1.19) | < 0.01 | 1.21 | (1.15-1.28) | < 0.01 | 1.25 | (1.16-1.33) | < 0.01 |
| 2 | Admission NIHSS | 1.16 | (1.12-1.20) | < 0.01 | 1.15 | (1.11-1.18) | < 0.01 | 1.21 | (1.15-1.28) | < 0.01 | 1.24 | (1.16-1.33) | < 0.01 |
|  | Time to assessment | 0.89 | (0.76-1.04) | 0.13 | 0.88 | (0.75-1.03) | 0.11 | 0.96 | (0.79-1.17) | 0.67 | 0.93 | (0.73-1.19) | 0.58 |
| 3 | Admission NIHSS | 1.11 | (1.05-1.17) | < 0.01 | 1.08 | (1.02-1.14) | < 0.01 | 1.23 | (1.13-1.33) | < 0.01 | 1.28 | (1.16-1.41) | < 0.01 |
|  | Time to assessment | 0.68 | (0.48-0.94) | 0.02 | 0.62 | (0.44-0.89) | < 0.01 | 1.04 | (0.70-1.57) | 0.84 | 1.09 | (0.71-1.68) | 0.68 |
|  | Interaction | 1.02 | (1.00-1.04) | 0.04 | 1.03 | (1.01-1.05) | 0.01 | 0.99 | (0.97-1.02) | 0.63 | 0.99 | (0.97-1.01) | 0.42 |
| 4 | Continuous centiles | 1.06 | (1.04-1.07) | < 0.01 | 1.05 | (1.04-1.07) | < 0.01 | 1.06 | (1.04-1.08) | < 0.01 | 1.07 | (1.05-1.10) | < 0.01 |

\* We adjusted for age, sex, pre-stroke mRS, medical history including cerebral infarction, subarachnoid haemorrhage, myocardial infarction, angina, peripheral vascular disease, hypertension, diabetes, paroxysmal atrial fibrillation, chronic atrial fibrillation in the model.

### Appendix D Comparisons of model performances in prediction mortality

We assessed the model performance similarly in predicting mortality in Table 5. For AIS patients, Model 3 achieved the best balance of fit with a Pseudo-R squared of 0.226, the BIC of 478, and an AUC of 0.850, indicating that it provided a slightly better model performance than others. For ICH patients, the time-incorporated methods did not improve the performance of the prognostic model. The adjusted models improved discrimination power but not model fit.

**Table 5.** Comparisons of model performances.

| Models | Acute ischaemic stroke |  |  |  |  |  | Intracerebral haemorrhage |  |  |  |  |  |
| --- | --- | --- | --- | --- | --- | --- | --- | --- | --- | --- | --- | --- |
|  | Unadjusted model |  |  | Adjusted model* |  |  | Unadjusted model |  |  | Adjusted model* |  |  |
|  | Pseudo-R<sup>2</sup> | BI<br>C | AUC | Pseudo-R<sup>2</sup> | BI<br>C | AUC | Pseudo-R<sup>2</sup> | BI<br>C | AUC | Pseudo-R<sup>2</sup> | BIC | AUC |
| 1 | 0.211 | 471 | 0.835 | 0.246 | 544 | 0.851 | 0.393 | 203 | 0.850 | 0.543 | 233 | 0.905 |
| 2 | 0.216 | 476 | 0.848 | 0.252 | 549 | 0.863 | 0.393 | 208 | 0.850 | 0.545 | 239 | 0.905 |
| 3 | 0.226 | 478 | 0.850 | 0.266 | 549 | 0.868 | 0.394 | 213 | 0.851 | 0.547 | 244 | 0.905 |
| 4 | 0.167 | 493 | 0.815 | 0.207 | 564 | 0.830 | 0.343 | 214 | 0.839 | 0.504 | 244 | 0.895 |

\* We adjusted for age, sex, pre-stroke mRS, medical history including cerebral infarction, subarachnoid haemorrhage, myocardial infarction, angina, peripheral vascular disease, hypertension, diabetes, paroxysmal atrial fibrillation, chronic atrial fibrillation in the model.

### Appendix E Test of proportional odds assumption for ordinal logistic regression

In the ordinal logistic regression model, the proportional odds assumption requires that the effect of an explanatory variable—in this case, the “estimated continuous centile”—is consistent across all levels of the outcome variable. This implies a common slope (β) for each cumulative logit, meaning the relationship between the predictor and the odds of being in a higher category of the outcome is assumed to be the same for all outcome thresholds. To test this assumption, we conducted the Brant test, which evaluates whether the effect of the explanatory variable differs significantly across cumulative logits. The null hypothesis (H0) posits that the slopes do not differ, indicating the proportional odds assumption is valid. If the p-value is large, we fail to reject H0, suggesting the proportional odds assumption holds. Conversely, a small p-value would lead to rejecting H0, indicating that the assumption does not hold, and the ordinal model may not be appropriate.

Table 6 presents the results of the Brant test for the continuous centile model. For patients with ischaemic stroke (AIS), the test statistic was 31.02, with a p-value of 0, indicating a violation of the proportional odds assumption. This suggests that the effect of the estimated continuous centile differs across cumulative logits in this group, making the ordinal model less suitable for AIS patients. For intracerebral haemorrhage (ICH) patients, however, the test statistic was 3.99 with a p-value of 0.55, suggesting no significant difference in slopes across logits. Thus, the proportional odds assumption is reasonable for ICH patients, and the ordinal logistic model is appropriate for this cohort.

**Table 6.** The results of the Brant test of the continuous centile model.

| Model | Test statistics | p-values |
| --- | --- | --- |
| AIS | 31.02 | 0 |
| ICH | 3.99 | 0.55 |

### Appendix F Model comparisons in ordinal logistic regression

**Table 7.** Model comparisons in ordinal logistic regression.

| Models | BIC |  |  |  |
| --- | --- | --- | --- | --- |
|  | Acute ischaemic stroke |  | Intracerebral haemorrhage |  |
|  | Unadjusted | Adjusted* | Unadjusted | Adjusted* |
| 1 | 6653 | 6463 | 799 | 822 |
| 2 | 6651 | 6462 | 803 | 824 |
| 3 | 6643 | 6453 | 807 | 828 |
| 4 | 6687 | 6522 | 833 | 858 |

\* We adjusted for age, sex, pre-stroke mRS, medical history including cerebral infarction, subarachnoid haemorrhage, myocardial infarction, angina, peripheral vascular disease, hypertension, diabetes, paroxysmal atrial fibrillation, chronic atrial fibrillation in the model.

### Appendix G Sensitivity analysis: Different ways to dichotomise mRS (0 to 1 vs 2 to 6)

**Table 8.** Model comparisons in logistic regression with mRS dichotomised as 0 to 1 vs 2 to 6.

| Models | Acute ischaemic stroke |  |  |  |  |  | Intracerebral haemorrhage |  |  |  |  |  |
| --- | --- | --- | --- | --- | --- | --- | --- | --- | --- | --- | --- | --- |
|  | Unadjusted model |  |  | Adjusted model* |  |  | Unadjusted model |  |  | Adjusted model* |  |  |
|  | Pseudo-R<sup>2</sup> | BIC | AUC | Pseudo-R<sup>2</sup> | BIC | AUC | Pseudo-R<sup>2</sup> | BIC | AUC | Pseudo-R<sup>2</sup> | BIC | AUC |
| 1 | 0.268 | 2359 | 0.764 | 0.390 | 2206 | 0.816 | 0.344 | 121 | 0.880 | 0.532 | 161 | 0.943 |
| 2 | 0.271 | 2361 | 0.764 | 0.393 | 2208 | 0.816 | 0.347 | 126 | 0.884 | 0.532 | 166 | 0.944 |
| 3 | 0.286 | 2341 | 0.767 | 0.402 | 2195 | 0.818 | 0.351 | 131 | 0.885 | 0.533 | 172 | 0.944 |
| 4 | 0.270 | 2357 | 0.763 | 0.390 | 2206 | 0.816 | 0.246 | 134 | 0.832 | 0.445 | 173 | 0.913 |

\* We adjusted for age, sex, pre-stroke mRS, medical history including cerebral infarction, subarachnoid haemorrhage, myocardial infarction, angina, peripheral vascular disease, hypertension, diabetes, paroxysmal atrial fibrillation, chronic atrial fibrillation in the model.

### Appendix H ROC Curves

**Figure 3.**
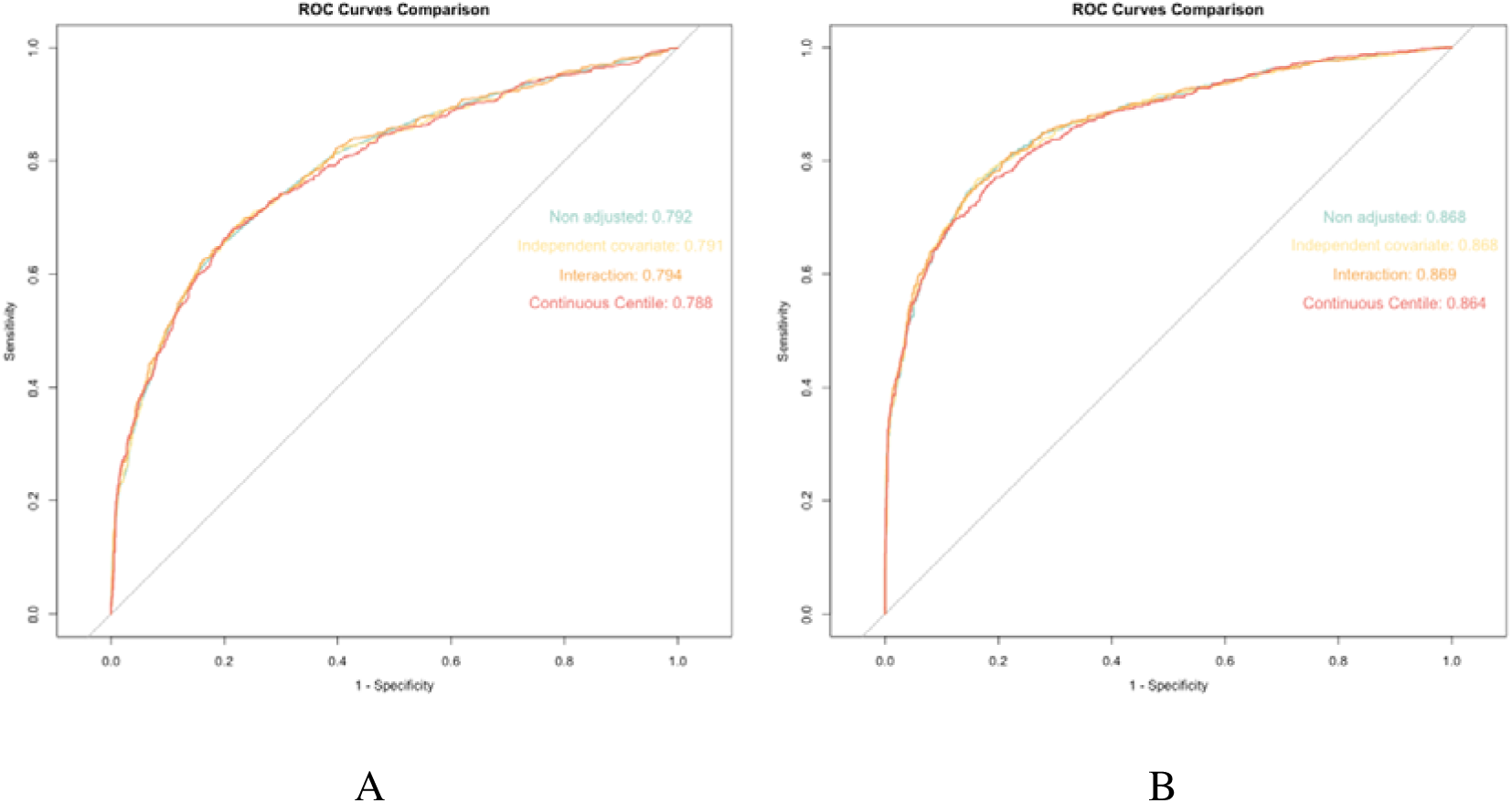
ROC curves with AUC when predicting functional dependency for AIS patients: A for unadjusted model and B for adjusted model

**Figure 4.**
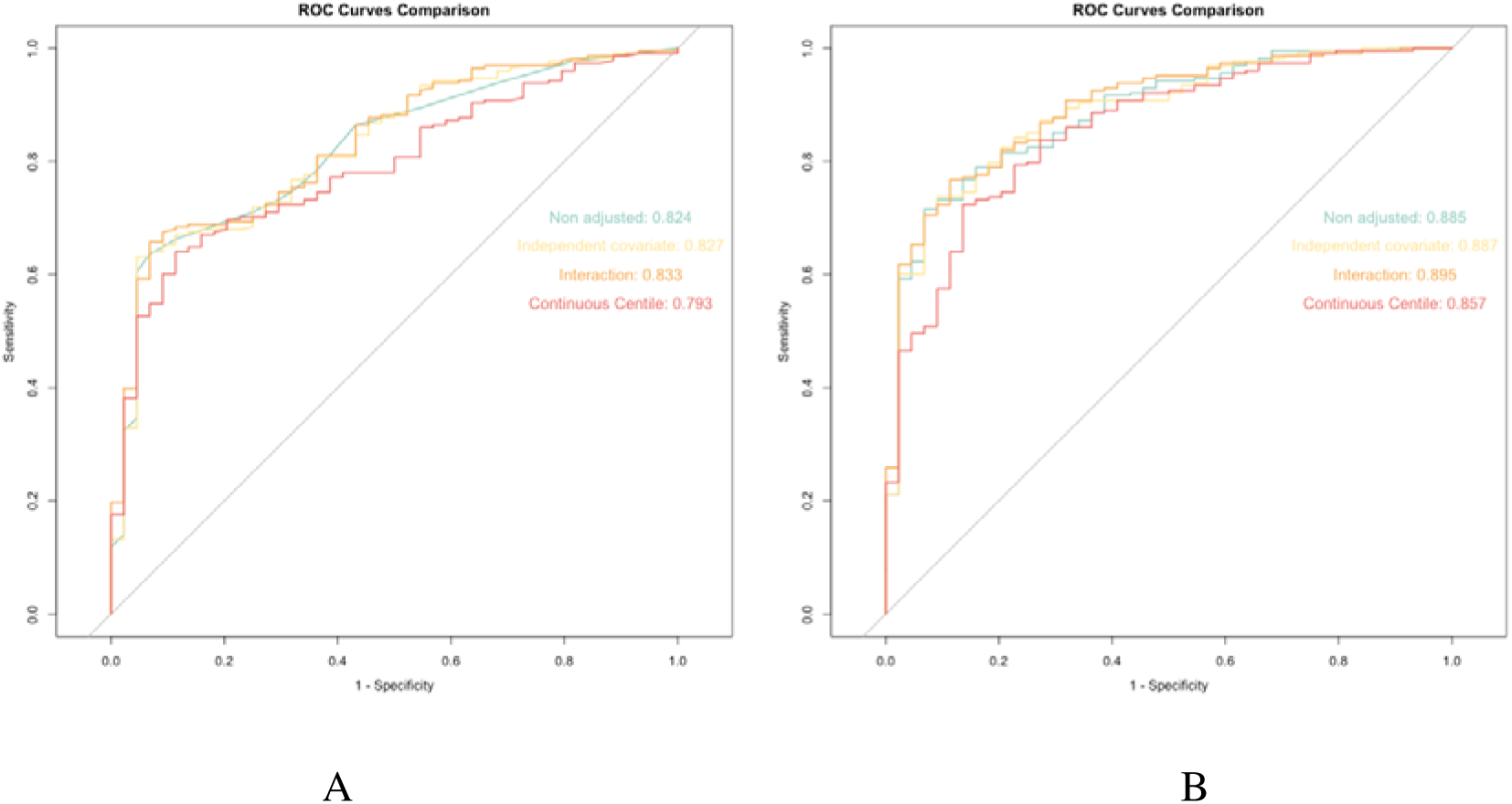
ROC curves with AUC when predicting functional dependency for ICH patients: A for unadjusted model and B for adjusted model

**Figure 5.**
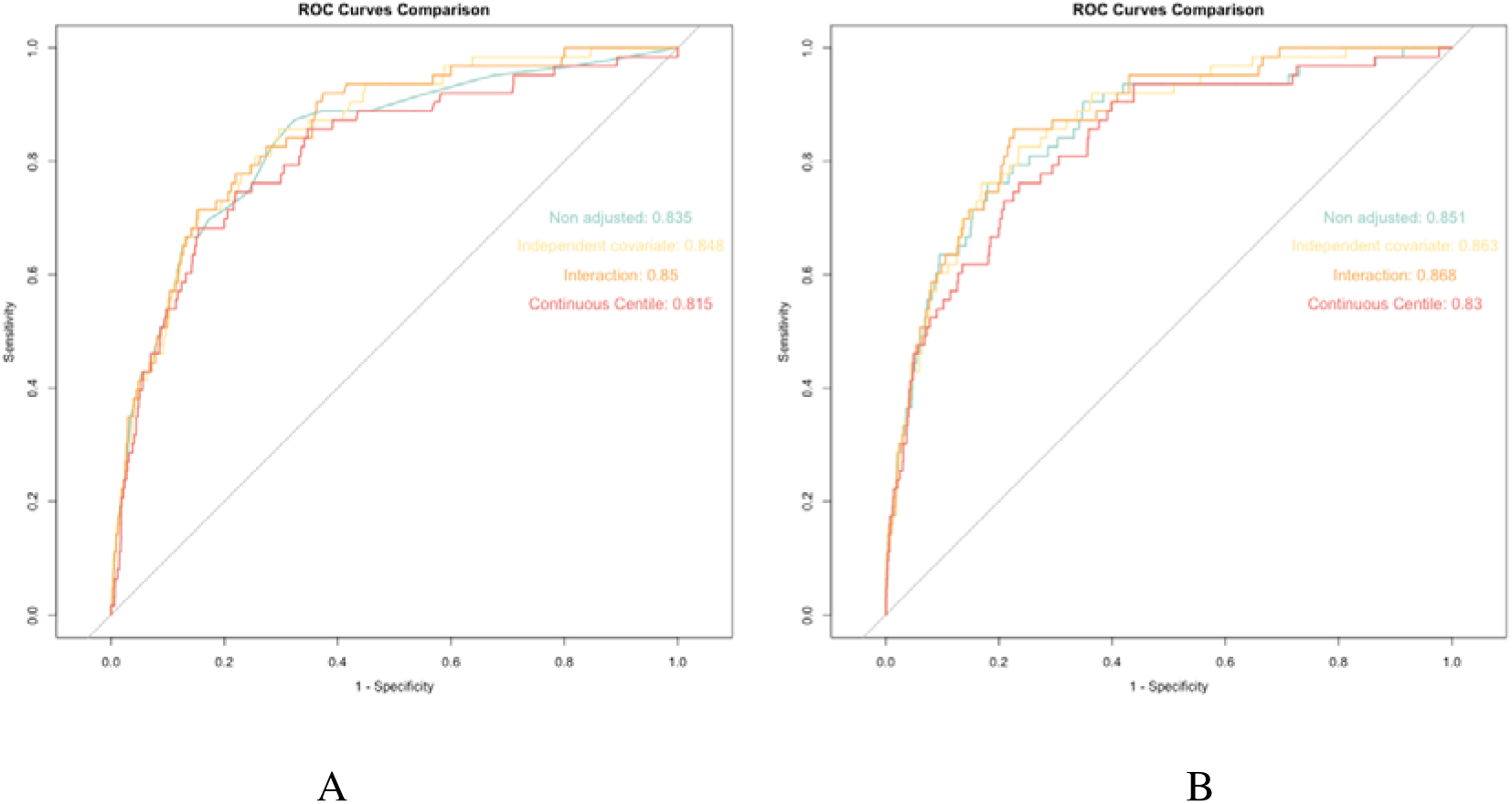
ROC curves with AUC when predicting mortality for AIS patients: A for unadjusted model and B for adjusted model

**Figure 6.**
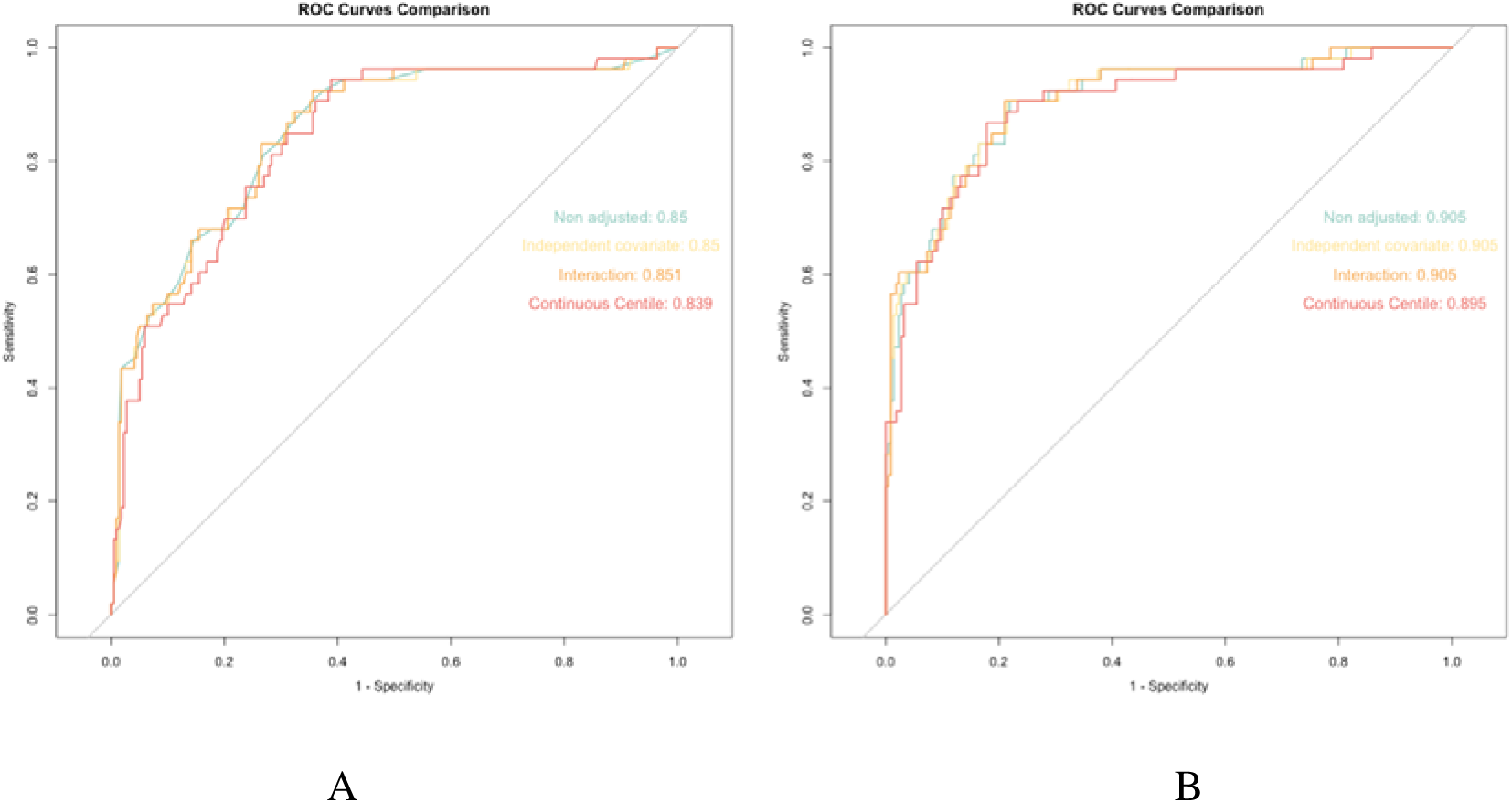
ROC curves with AUC when predicting mortality for ICH patients: A for unadjusted model and B for adjusted model

